# Predicting overall survival of NSCLC patients with clinical, radiomics and deep learning features

**DOI:** 10.1101/2025.06.13.25329594

**Authors:** Hemalatha Kanakarajan, Jikai Zhou, Aiara Lobo Gomes, Petros Kalendralis, Wenjie Liang, Fariba Tohidinezhad, Andre Dekker, Wouter De Baene, Margriet Sitskoorn

## Abstract

**Background and purpose:** Accurate estimation of Overall Survival (OS) in Non-Small Cell Lung Cancer (NSCLC) patients provides critical insights for treatment planning. While previous studies have shown that radiomics or Deep Learning (DL) features improved prediction accuracy, this study aimed to evaluate whether a model that integrates clinical, radiomics, DL, and dosimetric features outperforms other models developed with only a subset of these features.

**Materials and methods:** We collected pre-treatment lung CT scans and clinical data for 225 NSCLC patients from the Maastro Clinic: 180 for training and 45 for testing. Radiomics features were extracted using the Python radiomics feature extractor, and DL features were obtained using a 3D ResNet model. An ensemble model comprising XGB and NN classifiers was developed using: (1) clinical features only; (2) clinical and radiomics features; (3) clinical and DL features; and (4) clinical, radiomics, and DL features. The performance metrics were evaluated for the test and K-fold cross-validation data sets.

**Results:** The prediction model utilizing only clinical variables provided an Area Under the Receiver Operating Characteristic Curve (AUC) of 0.64 and a test accuracy of 77.55%. The best performance came from combining clinical, radiomics, and DL features (AUC: 0.84, accuracy: 85.71%). The prediction improvement of this model was statistically significant compared to models trained with clinical features alone or with a combination of clinical and radiomics features.

**Conclusion:** Integrating radiomics and DL features with clinical characteristics improved the prediction of OS after radiotherapy for NSCLC patients. The increased accuracy of our integrated model enables personalized, risk-based treatment planning, guiding clinicians toward more effective interventions, improved patient outcomes and enhanced quality of life.

## Introduction

Lung cancer is the most commonly diagnosed malignancy worldwide and remains the leading cause of cancer-related mortality. In 2022, approximately 2.5 million new cases and 1.8 million deaths were reported globally [1]. Based on histopathological classification, lung cancer is broadly categorized into Small Cell Lung Cancer (SCLC) and Non-Small Cell Lung Cancer (NSCLC), with NSCLC accounting for 85% to 90% of all cases [2]. Approximately one-quarter of NSCLC patients are diagnosed with a locally advanced stage, and the majority of these cases are considered inoperable [3].

Patients with unresectable locally advanced NSCLC display substantial heterogeneity, resulting in marked variability in Overall Survival (OS) [40]. OS is defined as the time from the initiation of treatment to patient death, irrespective of the cause. In the TNM staging system, NSCLC is categorized from stage I to IV based on tumor size, regional lymph node involvement, and the presence of distant metastases. Stage III disease, commonly referred to as locally advanced NSCLC, represents a heterogeneous group and is the central focus of the present study. This stage is further subdivided into IIIA, IIIB, and IIIC, each associated with distinct prognostic implications. The 5-year OS rates for patients with stage IIIA, IIIB, and IIIC are approximately 36%, 26%, and 13%, respectively [4], with smoking status significantly influencing the prognosis. In a retrospective study of 2,295 NSCLC patients, the median OS was 13.0 months for current smokers, 16.1 months for former smokers, and 29.0 months for never-smokers [45]. Moreover, additional factors such as tumor size, comorbidities, and treatment strategies (including radiotherapy volume, concurrent versus sequential chemoradiotherapy, and use of consolidation immunotherapy) have all been associated with survival differences [46–48].

OS remains a universally accepted and clinically meaningful endpoint in oncology. In NSCLC, where long-term prognosis is often poor, despite recent therapeutic advancements, accurate OS prediction is of paramount importance. First, it enables clinicians to stratify patients more precisely based on prognostic risk, thereby facilitating individualized treatment planning. Second, it supports more efficient allocation of healthcare resources by identifying patients who are most likely to benefit from intensified therapies or enrollment in clinical trials. Third, it offers more reliable prognostic information to patients and their families, aiding in shared decision-making prior to treatment and enhancing psychological readiness for care [49]. These considerations highlight the substantial clinical value of developing machine learning models capable of accurately predicting OS and constitute the primary motivation for this study.

Prior research has explored various machine learning approaches to enhance OS prediction in NSCLC patients. Siah et al. (2019) developed machine-learning models to predict OS in NSCLC patients, achieving a Concordance Index (C-index) of 0.73 (95% CI, 0.72–0.74) [5]. They identified biomarker status—specifically PD-L1 expression, EGFR mutation, and ALK rearrangement—as the strongest predictor for OS. However, the limited predictive value of single biomarkers emphasized the need for more comprehensive approaches to improve prediction accuracy [5]. Maria et al. (2024) also emphasized the critical need to develop reliable and accurate tools to predict OS before treatment initiation [50], a perspective shared by other researchers [51]. In this context, radiomics and deep learning have proven to be powerful, complementary tools that leverage advanced imaging analysis and artificial intelligence to enhance OS prediction in NSCLC patients through more accurate, non-invasive, and personalized assessments [12, 37, 52, 53, 54].

Radiomics is a research field that extracts quantifiable data, such as patterns, shapes, and intensity levels, from medical images [6]. These data are used to describe and better understand tumors or other irregularities seen in the images, helping doctors and researchers in diagnosis and treatment planning. The radiomics-based approach has been successfully applied for different endpoints in lung cancer, including overall and progression-free survival [7,8,10]. Recent studies have explored radiomics-based approaches for predicting OS in NSCLC. Sun et al. (2018) applied machine learning to radiomic features in 283 patients and achieved a C-index of 0.68 (95% CI, 0.62–0.74) for OS prediction [7]. Liu et al. (2021) developed a CT-based radiomics model for patients receiving nivolumab, reporting an average AUC of 0.61 ± 0.08 for OS and a C-index of ∼0.79 [8]. The results of these studies highlight the applicability of radiomics features in predicting OS and subsequently improving the prognostic accuracy of NSCLC patients.

Deep learning, a groundbreaking advancement in artificial intelligence, has transformed numerous fields, including image processing, computer vision, and natural language processing [9]. Deep learning models have also been used to extract imaging features for cancer treatment [24, 25, 26]. In medical imaging, deep learning methods are used to learn the hierarchical representations of the data. The information extracted by the deep learning models from the images can help in predicting treatment outcomes [10, 19, 20, 21, 22, 27, 28, 29]. Recent studies have explored the application of deep learning in predicting OS for NSCLC patients. Deep learning models using CT scans have shown significant potential for predicting survival and cancer-specific outcomes in NSCLC patients [22]. Hosny et al. (2018) demonstrated that a 3D CNN could predict OS in patients treated with radiotherapy or surgery, achieving AUCs of 0.70 (95% CI, 0.63–0.78) and 0.71 (95% CI, 0.60–0.82), respectively— outperforming hand-crafted radiomics and clinical models [19]. Afshar et al. (2020) developed the deep learning–based radiomics model, reporting a C-index of 0.68 for OS prediction, notably higher than that of traditional radiomics (≈0.51) [20]. Peng et al. (2024) developed a 3D ResNet50-based model using contrast-enhanced CT to estimate response to concurrent chemoradiotherapy in NSCLC, achieving an AUC of 0.84 in the validation cohort and showing a significant association with OS [21]. Similarly, Xu et al. (2019) integrated serial CT scans using a combined CNN–RNN framework in stage III NSCLC patients, achieving an AUC of 0.74 for two-year OS prediction [10]. Collectively, these studies demonstrate that deep learning models can match or exceed the performance of hand-crafted radiomics in predicting the clinical outcomes.

Integrating diverse data sources has also shown promise. Lai et al. (2020) demonstrated that integrating clinical data into deep learning networks helps develop high accuracy OS prediction models for NSCLC patients [11]. Similarly, combining clinical and radiomics features extracted from CT images has improved prediction accuracy [12, 13, 14, 15]. Caii et al. (2024) explored integrating radiomics and deep learning features to predict OS in NSCLC patients treated with immunotherapy [23]. Hou et al., (2024) further demonstrated the significance of dose distribution maps by combining them with deep learning features to predict the Progression-Free Survival (PFS) of NSCLC patients [31].

To the best of our knowledge, no study has yet integrated hand crafted radiomics and deep learning features with clinical features and dose information to develop machine learning algorithms to predict OS for NSCLC patients. A model trained with all these combined features might predict OS with higher accuracy than models trained with partial combinations of these features. This study aimed to develop a prediction model that combines the radiomics and deep learning features from the images with the clinical and dose information to predict 12-month OS classification for NSCLC patients.

Predicting OS with high accuracy provides a more comprehensive understanding of treatment response prior to treatment initiation and could lead to more personalized and effective interventions that improve treatment outcomes, prolong patient survival, and enhance quality of life.

## Methods

### Data collection

We retrospectively collected clinical data for 296 NSCLC patients from the Maastro Clinic in Maastricht, The Netherlands. All patients were aged 18 or older. This study was approved by the Institutional Review Board of Maastro Clinic (P0617) and Ethics Committee of Maastricht University Medical Centre (METC 2023-0377). All patients underwent definitive radiotherapy (54–66 Gy) between 2015 and 2022. Some patients received concurrent or sequential chemotherapy in addition to radiotherapy, and others received consolidation immunotherapy with durvalumab. Patients with lung surgery or chest radiotherapy history were excluded, as such treatments may alter tumor morphology, density, or imaging texture, potentially affecting the extraction of radiomics and deep learning features. None of the selected patients had any other primary tumor. Although locally advanced NSCLC is conventionally defined as stage III disease, we included both unresectable stage II and stage III patients in this study. This decision was based on the clinical observation that unresectable stage II and III patients often receive comparable treatment regimens, including definitive radiotherapy and systemic therapies [58, 59, 60]. Additionally, including stage II patients allowed for an increased sample size, thereby enhancing the statistical power and generalizability of our predictive model.

For these 296 patients, we collected the planning CT images taken before radiotherapy. CT images were acquired using a Siemens scanner (software version 4.2.7.0). Scanning parameters included a tube current of 60 mA, a slice thickness of 3 mm, and a reconstruction diameter of 650 mm. Images were reconstructed using the Qr40f convolution kernel. Pixel spacing was 1.27 mm × 1.27 mm, and images were stored in 16-bit format with Hounsfield Unit (HU) scaling (rescale slope = 1, intercept = – 1000). The acquisition covered the thoracic region, and all patients were positioned Head-First Supine (HFS). For all patients, the segmentations of the tumors were obtained through manual delineation by expert radiation oncologists from the Maastro Clinic.

From the initial sample of 296 patients, all patients whose OS status was not known at 12 months from radiotherapy were excluded (n = 46). We also excluded 25 patients for whom we had issues in extracting the radiomics features from the Lung gross tumor volume segmentation (either because the segmentation was missing or more than one segmentation was present). This resulted in a total of 225 patients. For patients with more than one planning CT scan, we took the CT scan that was closest to the radiotherapy date.

### Clinical features

The list of clinical factors that we collected from the Maastro clinic were age at radiotherapy, gender, BMI, T classification, N classification, NSCLC stage, histological subtype, laterality, location, World Health Organisation Performance Status (WHOPS), smoking status, presence of diabetes, presence of hypertension, presence of cardiac history, presence of cardiac surgery, presence of vascular issues, presence of cerebra vascular issues, presence of respiratory issues, presence of neoplasm, presence of kidney diseases, presence of liver diseases, creatinine level, CKD-EPI (Chronic Kidney Disease Epidemiology Collaboration) level, CRP (C-reactive protein) level, FEV₁ (Forced Expiratory Volume in 1 second), DLCO (Diffusing Capacity of the Lungs for Carbon Monoxide), type of chemo sequence (concurrent/sequential), durvalumab administered indicator, and the OS indicator at the end of 12 months from beginning of radiotherapy. We extracted the planned radiotherapy dose from the RTDOSE DICOM files, which contain 3D dose distributions generated by the treatment planning system. To ensure we used the complete planned dose for each patient, we selected only files where the ’DoseSummationType’ was set to ’PLAN’—indicating the total dose from the full treatment plan. The pixel values in these files were converted to physical dose values in Gray (Gy). Rather than using the full 3D dose distribution, we derived each patient’s total dose from the 3D dose map and added this value to the clinical dataset. This approach ensures accurate dose extraction directly from the source data.

Missing numerical clinical values were imputed using the iterative imputer provided by the Python scikit learn package (https://scikit-learn.org/stable/modules/impute.html). This iterative imputer models each feature with missing values as a function of other features and uses that estimate for imputation. This is done in an iterated round-robin fashion. At each step, a feature column is designated as output y, and the other feature columns are treated as inputs X. A regressor fits on (X, y) for known y. The regressor is then used to predict the missing values of y. This is done for each missing feature in an iterative fashion and is repeated for 10 imputation rounds. One of the limitations of imputing missing values is that when there are too many missing values, the model may not learn a good predictor for that feature. However, the missing clinical values for each of the factors in our clinical data did not exceed 15% of the total number of records, which is well below levels shown to affect model performance. In fact, Junaid et al [55] demonstrated that Multiple Imputation by Chained Equations (a method similar to IterativeImputer) remains effective, yielding results comparable to complete data, with minimal bias and variance distortion even with up to 50% of missing data. We imputed missing categorical values using probabilistic imputation and applied one-hot encoding [32].

### Radiomics features

The segment-based radiomics features were extracted from the radiotherapy treatment planning CT scans using the radiomics feature extractor of the Python Pyradiomics package. We extracted the radiomics features from the Lung gross tumor volume segmentation made by the expert radiation oncologists at Maastro clinic. The seven groups of features extracted from the Region Of Interest (ROI) of the tumor segmentations were shape-based features (14 features), first-order features (18), Gray Level Cooccurrence Matrix (GLCM) features (24), Gray Level Dependence Matrix (GLDM) features (14), Gray Level Run Length Matrix (GLRLM) features (16), Gray Level Size Zone Matrix (GLSZM) features (16), and Neighbouring Gray Tone Difference Matrix (NGTDM) features (5). The resulting 107 radiomics features were considered in this study. The list of radiomics features extracted is listed in the appendix. The definitions of these radiomics features are given in the Pyradiomics documentation (https://pyradiomics.readthedocs.io/en/latest/features.html). The IBSI (Image Biomarker Standardisation Initiative) has standardized the guidelines for extracting the radiomics features from medical images. PyRadiomics has incorporated most of the IBSI recommendations. The minor deviations from this standard are justified in the documentation (https://pyradiomics.readthedocs.io/en/latest/faq.html).

### Deep learning features

We extracted deep learning features from the CT scans using a pre-trained 3D ResNet-18 model [41]. The 3D ResNet-18 model is found to have superior performance when compared to other models for classification tasks [41]. The pre-processing steps included normalizing the pixel intensities between 0 and 1 and ensuring all images are in uniform size by cropping or padding. We extracted the deep features from the output of the GlobalAveragePooling3D layer, which is the second-to-last layer in the model. A 3D convolution was applied to the training data. This convolutional layer was designed with a large kernel size of (3, 7, 7), a stride of (1, 2, 2) for down sampling, and padding of (1, 3, 3) to control the spatial dimensions of the output. The 512 deep learning features extracted using this fine-tuned 3D ResNet model were combined with clinical and radiomics features to form a combined Python dataframe. The complete process of pre-processing and feature extraction is summarized in Figure 1.

**Figure 1:**
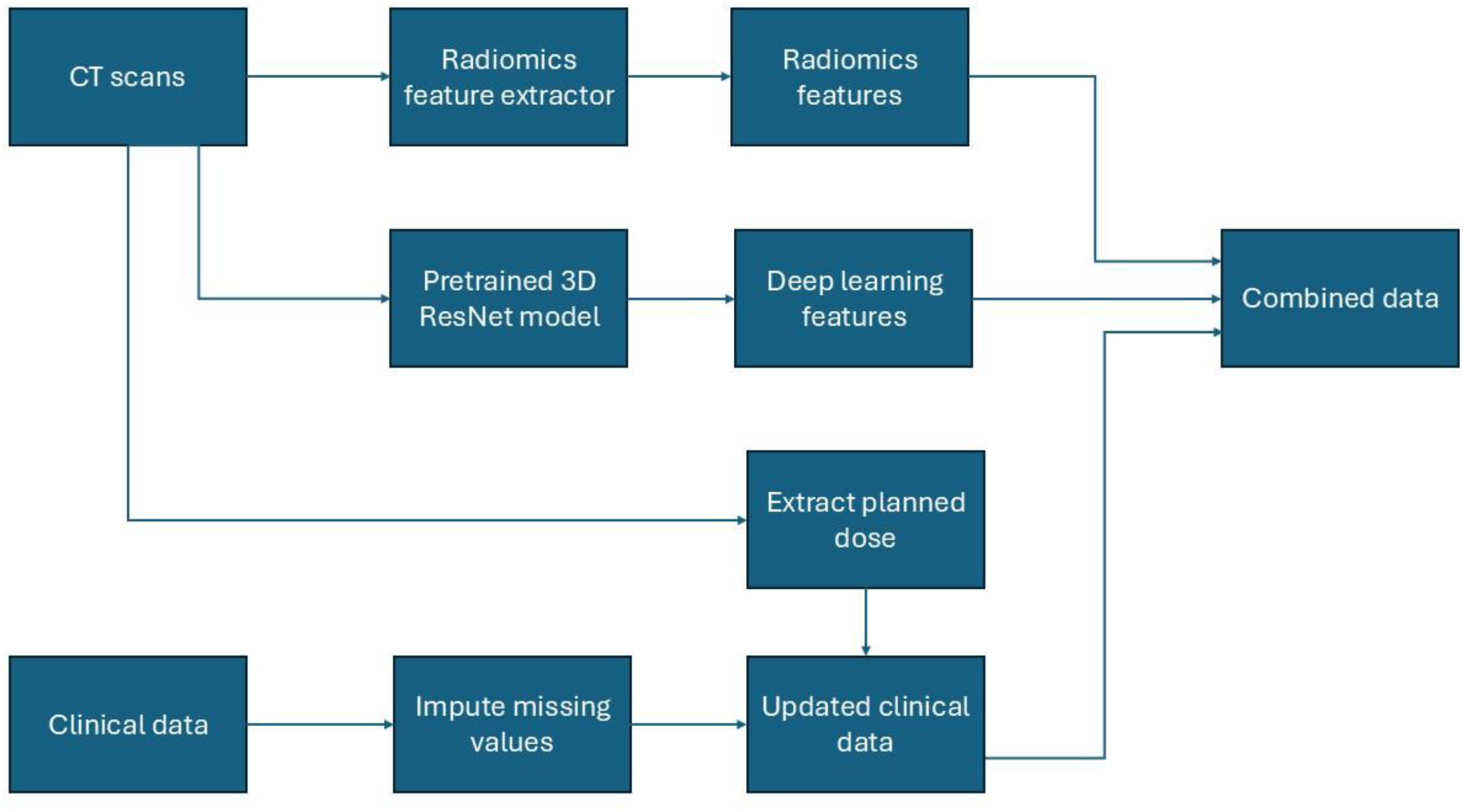
The process of pre-processing, feature extraction, and combining the data.

### Model training

We standardized the combined features using the StandardScalar of the Python scikit-learn library (https://scikit-learn.org/stable/modules/generated/sklearn.preprocessing.StandardScaler.html).

The features with low variance (<0.1) were determined and excluded from the combined dataset to improve prediction accuracy [42]. We applied feature selection using Recursive Feature Elimination (RFE) with a Random Forest Classifier. RFE iteratively removes the least important features and has the potential to improve model performance and reduce overfitting. After RFE, we included only the top 50 most important features. This number is based on the performance of the models in our trial runs with random counts and with the default count (which is half the number of original features). The data set was then divided into a training data set (80%) and a test data set (20%). The training data set contained 180 patients, and the test data set contained 45 patients. We balanced the training data set using SMOTE (oversampling) [33] and applying random under sampling using Python RandomUnderSampler (https://imbalanced-learn.org/stable/references/generated/imblearn.under_sampling.RandomUnderSampler.html). We did oversampling and then undersampling to resolve the class imbalance. The comprehensive analysis of Imani et al. [43] showed that for datasets which are balanced with SMOTE, XGBoost consistently achieved better classification performance when compared to the Random Forest algorithm. Also, the study of Fatima et al. [44] showed that XGBoost consistently achieved better performance for predictive modelling. So, we developed an XGBoost (XGB) classifier for OS classification. We also developed a Neural Network (NN) classifier and compared the performance of the XGB classifier with the NN classifier. We trained the XGB and NN classifiers separately using the training data set. We fine-tuned the hyperparameters of the models to establish the values for the best performance. For XGB, the values are n_estimators=100, learning_rate=0.1, max_depth=4, and scale_pos_weight=1.

We designed our NN classifier using two linear layers with a ReLU (Rectified Linear Unit) activation function in between. ReLU introduces non-linearity into the model by setting all negative values to zero while keeping positive values unchanged, helping the network learn complex patterns more effectively [34, 35]. For training, we used the BCEWithLogitsLoss loss function (https://pytorch.org/docs/stable/generated/torch.nn.BCEWithLogitsLoss.html), which conveniently combines binary cross-entropy loss with a sigmoid activation in a single step. This loss function simplifies implementation while maintaining accuracy and stability. We optimized the model using Adam, an adaptive learning rate optimizer that adjusts learning rates for individual parameters, enabling faster and more robust convergence. The initial learning rate was set to 0.001. To further improve training stability, we used a learning rate scheduler that decreased the learning rate every five epochs, scaling it to 80% of its current value. This gradual reduction helps the model make finer adjustments as training progresses, which often improves generalization and final performance. The model was trained for a total of 25 epochs, balancing computational efficiency with sufficient iterations for convergence.

Though XGB classifier achieved better performance for most of the evaluation metrics, NN classifier provided higher Recall when compared to XGB. To harness the advantages provided by both models, we created an ensemble model comprising both XGB classifier and NN classifier. Ensemble models have been shown to outperform individual models in classification tasks in the medical domain [56, 57]. The trained XGB and NN classifiers were combined to form the ensemble model which combines predictions from XGBoost and NN classifiers using a weighted average approach. The predictions are combined with weights of 0.75 for XGB and 0.25 for the NN classifier. The weights are based on the performance of the ensemble model in our trial runs with different weight combinations. We chose the weights which provided the best performance for all the evaluation metrics. The different ensemble models that we created were:

1. Ensemble model with clinical features only.
2. Ensemble model trained with clinical and deep learning features.
3. Ensemble model trained with clinical and radiomics features.
4. Ensemble model trained with clinical, radiomics and deep learning features.

### Model evaluation

The performance of the models was evaluated by measuring the following metrics: classification accuracy, precision, F1 score, recall, Area Under the receiver operating characteristic Curve (AUC), Matthews Correlation Coefficient (MCC), and specificity. The classification accuracy is the ratio of the number of correct classifications to the total number of samples. The precision is the ability of the classifier to identify positive samples correctly, and recall is the ability of the classifier to find all the positive samples. In other words, precision is the ratio of true positive classifications to the total number of positive classifications made by the model, while recall is the ratio of true positive classifications to the total number of actual positives in the dataset. The F1 score gives the balance between precision and recall. A Receiver Operating Characteristic (ROC) is created by plotting the true positive rate vs the false positive rate at various thresholds for defining a positive case. The AUC computes the area under the ROC curve. The MCC is a reliable metric for evaluating the performance of a binary classifier, especially in the presence of imbalanced datasets. The MCC takes into account all four quadrants of a confusion matrix and provides a balanced measure that considers both the positive and negative classes [36]. Specificity helps understand how well the model performs when predicting the negative class.

A K-fold cross-validation was applied on the model. The different values for K that we used during cross-validation were 5, 7, and 10. The average accuracy and other performance metrics across the different folds were calculated. The best performing model across the different cross validation folds was then tested with a held-out test data set. The accuracy and the other performance metrics were then calculated for the test data set. The pre-processing, feature extraction, model training, and evaluation were performed using Python (version 3.10).

### Comparison of models

To evaluate if there was a significant difference in the performance between the four models across the cross-validation data sets and to understand the significance of the performance differences, we statistically compared the performance metrics of the four models. We employed the Friedman test to analyze the significance of the difference in each of the performance metrics across all validation folds across all values of K. When the Friedman test showed a significant difference across the models, a pairwise comparison was done using Wilcoxon signed-rank test with a False Discovery Rate (FDR) correction using the Benjamini-Hochberg method.

## Results

### Patient characteristics

A total of 219 NSCLC patients were included in this study, among whom 165 (75.3%) were alive at 12 months and 54 (24.7%) had died. There were no statistically significant differences in most baseline characteristics between the 12-month survival and mortality groups, including age, gender, BMI, tumor stage, and comorbidities (all p > 0.05). However, performance status (WHO PS) and history of cardiac surgery showed significant differences. Patients with worse performance status (WHO PS 2–3) and those with prior cardiac surgery were more frequently observed in the 12-month mortality group (p < 0.05). In addition, the use of durvalumab immunotherapy was significantly higher in the survival group (24.2% vs. 5.6%, p = 0.0026), suggesting a potential survival benefit. Chemo-radiotherapy sequence showed a trend toward significance (p = 0.0542), with concurrent chemoradiation more common in survivors. However, this association should be interpreted with caution, as patients with better performance status were more likely to receive concurrent chemoradiation, indicating possible treatment selection bias. Detailed patient characteristics are summarized in Table 1.

**Table 1.** shows the characteristics of patients included in our study.

| Factor | 12-month survival | 12-month mortality | P Value* |
| --- | --- | --- | --- |
| Age (Year)<br>(mean±SD) | 67.34 ± 9.84 | 69.98 ± 8.67 | 0.0602 |
| Gender |  |  | 0.0822 |
| Female | 73 (44.2%) | 16 (29.6%) |  |
| Male | 92 (55.8%) | 38 (70.4%) |  |
| BMI | 25.93 ± 5.19 | 26.33 ± 6.05 | 0.7722 |
| T Stage |  |  | 0.8749 |
| 1 | 22 (13.3%) | 5 (9.3%) |  |
| 2 | 39 (23.6%) | 13 (24.1%) |  |
| 3 | 34 (20.6%) | 11 (20.4%) |  |
| 4 | 70 (42.4%) | 25 (46.3%) |  |
| N Stage |  |  | 0.8701 |
| 0 | 34 (20.6%) | 14 (25.9%) |  |
| 1 | 25 (15.2%) | 8 (14.8%) |  |
| 2 | 78 (47.3%) | 24 (44.4%) |  |
| 3 | 28 (17.0%) | 8 (14.8%) |  |
| Clinical stage |  |  | 0.3418 |
| IIA | 5 (3.0%) | 0 (0.0%) |  |
| IIB | 24 (14.5%) | 7 (13.0%) |  |
| IIIA | 61 (37.0%) | 27 (50.0%) |  |
| IIIB | 58 (35.2%) | 14 (25.9%) |  |
| IIIC | 17 (10.3%) | 6 (11.1%) |  |
| Laterality |  |  | 0.9188 |
| Left | 70 (42.4%) | 24 (44.4%) |  |
| Right | 95 (57.6%) | 30 (55.6%) |  |
| Location |  |  | 0.6622 |
| Bronchus/Hilum | 10 (6.1%) | 3 (5.6%) |  |
| Lower | 49 (29.7%) | 21 (38.9%) |  |
| Middle | 7 (4.2%) | 2 (3.7%) |  |
| Upper | 99 (60.0%) | 28 (51.9%) |  |
| WHOPS |  |  | <0.05 |
| 0 | 19 (11.5%) | 1 (1.9%) |  |
| 1 | 114 (69.1%) | 18 (33.3%) |  |
| 2 | 26 (15.8%) | 21 (38.9%) |  |
| 3 | 6 (3.6%) | 14 (25.9%) |  |
| Smoking |  |  | 0.6635 |
| Current | 45 (27.3%) | 12 (22.2%) |  |
| Former | 114 (69.1%) | 39 (72.2%) |  |
| Never | 6 (3.6%) | 3 (5.6%) |  |
| Diabetes |  |  | 0.7098 |
| No | 140 (84.8%) | 44 (81.5%) |  |
| Yes | 25 (15.2%) | 10 (18.5%) |  |
| Hypertension |  |  | 0.2000 |
| No | 67 (40.6%) | 16 (29.6%) |  |
| Yes | 98 (59.4%) | 38 (70.4%) |  |
| Cardiac History |  |  | 1.0000 |
| No | 102 (61.8%) | 33 (61.1%) |  |
| Yes | 63 (38.2%) | 21 (38.9%) |  |
| Cardiac Surgery |  |  | 0.0360 |
| No | 137 (83.0%) | 37 (68.5%) |  |
| Yes | 28 (17.0%) | 17 (31.5%) |  |
| Vascular issues |  |  | 0.0607 |
| No | 135 (81.8%) | 37 (68.5%) |  |
| Yes | 30 (18.2%) | 17 (31.5%) |  |
| Cerebro vascular issues |  |  | 0.6488 |
| No | 135 (81.8%) | 42 (77.8%) |  |
| Yes | 30 (18.2%) | 12 (22.2%) |  |
| Respiratory issues |  |  | 0.7781 |
| No | 79 (47.9%) | 24 (44.4%) |  |
| Yes | 86 (52.1%) | 30 (55.6%) |  |
| Neoplasm |  |  | 0.7990 |
| No | 133 (80.6%) | 42 (77.8%) |  |
| Yes | 32 (19.4%) | 12 (22.2%) |  |
| Kidney diseases |  |  | 0.1571 |
| No | 146 (88.5%) | 43 (79.6%) |  |
| Yes | 19 (11.5%) | 11 (20.4%) |  |
| Liver diseases |  |  | 0.1994 |
| No | 159 (96.4%) | 49 (90.7%) |  |
| Yes | 6 (3.6%) | 5 (9.3%) |  |
| Creatinine | 83.84 ± 32.26 | 97.37 ± 85.59 | 0.3974 |
| CKD-EPI | 74.50 ± 27.39 | 71.39 ± 28.42 | 0.3863 |
| CRP | 39.06 ± 59.48 | 35.43 ± 44.11 | 0.3789 |
| FEV1 | 73.47 ± 21.19 | 70.48 ± 20.94 | 0.3908 |
| DLCO | 63.65 ± 17.64 | 59.29 ± 20.38 | 0.2067 |
| Chemo Sequence |  |  | 0.0542 |
| Concurrent | 101 (61.2%) | 23 (42.6%) |  |
| Sequential | 27 (16.4%) | 14 (25.9%) |  |
| Without chemotherapy | 37 (22.4%) | 17 (31.5%) |  |
| Durvalumab immunotherapy |  |  | 0.0026 |
| No | 125 (75.8%) | 51 (94.4%) |  |
| Yes | 40 (24.2%) | 3 (5.6%) |  |
| Prescribed total dose(Gy) | 61.70 ± 2.85 | 61.62 ± 2.54 | 0.9176 |

*Abbreviations:* SD = standard deviation; BMI = body mass index; CKD-EPI = Chronic Kidney Disease Epidemiology Collaboration equation; CRP = C-reactive protein; FEV<sub>1</sub> = forced expiratory volume in 1 second; DLCO = diffusing capacity of the lung for carbon monoxide.
\*P-values were calculated using the Chi-square test or Fisher's exact test for categorical variables, and Welch's t-test or Mann-Whitney U test for continuous variables, as appropriate. We added the P-values to this table to show the statistical comparison between the two groups on every factor.

The average performance metrics for the four ensemble models across the cross-validation datasets are shown in Table 2. In bold is the value of the best score for the corresponding performance metric.

**Table 2:** Average performance of the models along with their 95% confidence intervals on the validation datasets.

| Model | Accuracy (%) | Precision (%) | Recall (%) | F1 score (%) | AUC | MCC | Specificity (%) |
| --- | --- | --- | --- | --- | --- | --- | --- |
| Clinical features only | 78.65<br>(75.91, 81.4) | 71.64<br>(66.5, 76.77) | 80.07<br>(77.27, 82.87) | 74.74<br>(71.47, 78.00) | 0.79<br>(0.77, 0.82) | 0.57<br>(0.52, 0.62) | 78.30<br>(74.95, 81.65) |
| Clinical and deep learning features | 81.23<br>(76.87, 85.58) | 74.05<br>(68.75, 79.36) | 85.55<br>(80.04, 91.06) | 78.75<br>(74.31, 83.18) | 0.82<br>(0.78, 0.87) | 0.63<br>(0.54, 0.72) | 79.23<br>(74.42, 84.14) |
| Clinical and radiomics features | 77.67<br>(75.34, 80.00) | 68.95<br>(65.32, 72.58) | 80.00<br>(76.06, 84.52) | 73.55<br>(70.78, 76.32) | 0.78<br>(0.76, 0.80) | 0.55<br>(0.51, 0.60) | 76.16<br>(73.41, 78.91) |
| Clinical, radiomics and deep learning features | **82.61**<br>**(80.10, 85.12)** | **74.75**<br>**(70.86, 78.83)** | **86.62**<br>**(82.44, 90.78)** | **79.62**<br>**(76.96, 82.23)** | **0.84**<br>**(0.81, 0.87)** | **0.65**<br>**(0.60, 0.71)** | **80.84**<br>**(78.09, 83.60)** |

The accuracy of the model trained with clinical features only was 78.65%. The model trained with the combination of clinical and deep learning features had an accuracy of 81.23%. The accuracy of the model trained with the combination of clinical and radiomics features was 77.67%. However, the model with the highest prediction accuracy of 82.61% was the model trained with the combination of clinical, radiomics, and deep learning features. This model also achieved the highest precision of 74.75%, Recall of 86.62%, F1 score of 79.62%, MCC of 0.65, specificity of 80.84% and the best AUC of 0.84. The model trained with the combination of all the features had the best score for all performance metrics.

The performance metrics for the four ensemble models for the test data set are shown in Table 3. In bold is the value of the best score for the corresponding performance metric. The performance metrics of the separate XGB and NN classifiers for the test data set are added to the Appendix.

**Table 3:**
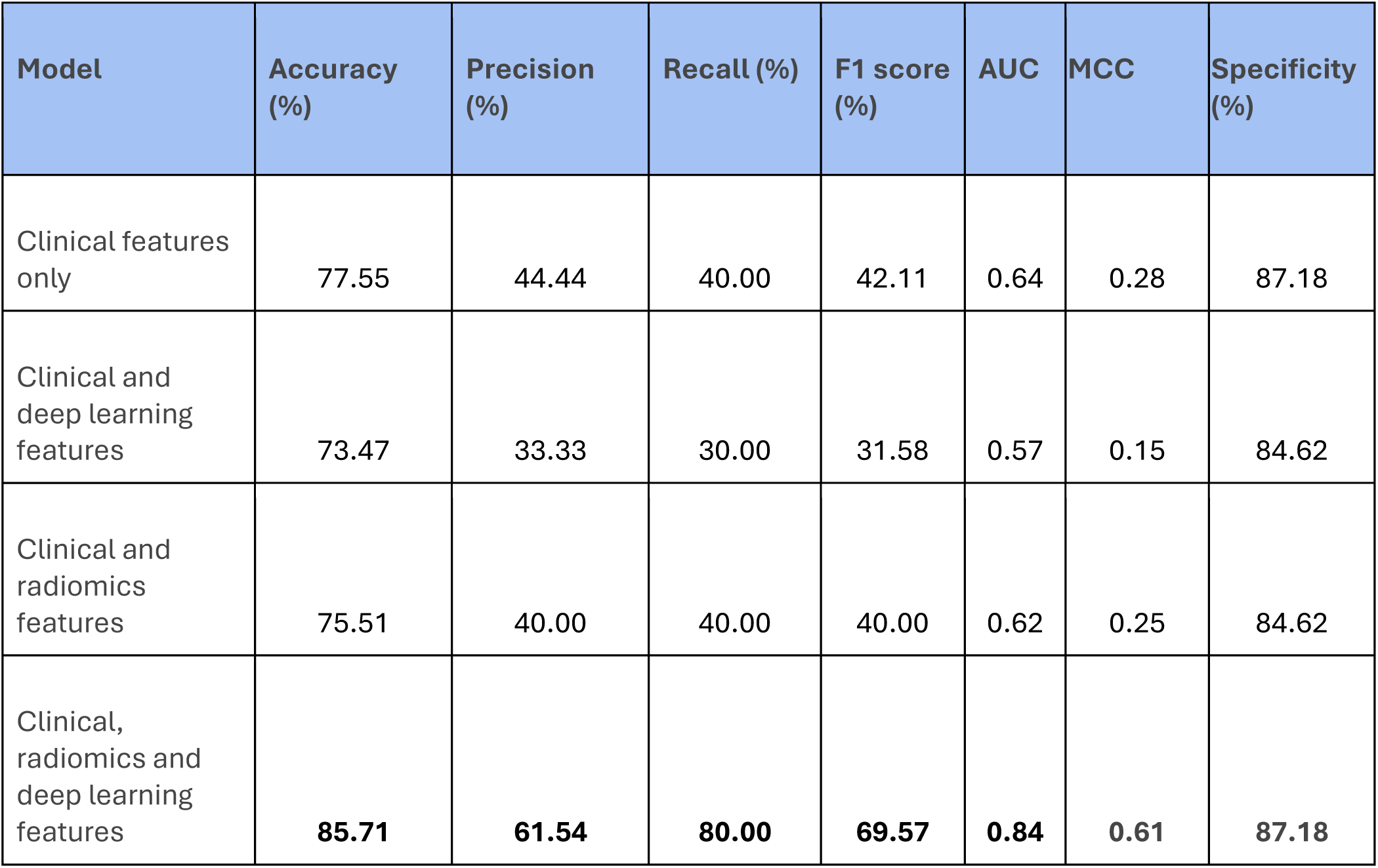
Performance of the models for the test dataset.

| Model | Accuracy (%) | Precision (%) | Recall (%) | F1 score (%) | AUC | MCC | Specificity (%) |
| --- | --- | --- | --- | --- | --- | --- | --- |
| Clinical features only | 77.55 | 44.44 | 40.00 | 42.11 | 0.64 | 0.28 | 87.18 |
| Clinical and deep learning features | 73.47 | 33.33 | 30.00 | 31.58 | 0.57 | 0.15 | 84.62 |
| Clinical and radiomics features | 75.51 | 40.00 | 40.00 | 40.00 | 0.62 | 0.25 | 84.62 |
| Clinical, radiomics and deep learning features | **85.71** | **61.54** | **80.00** | **69.57** | **0.84** | **0.61** | **87.18** |

Similar to the performance metrics for the cross validation data set, the model trained with the combination of all the features had the best score for all performance metrics for the test data set. The ROC curve of the model trained with the combination of clinical, radiomics, and deep learning features is shown in Figure 3. The ROC curves of the other three models were added to the Appendix.

**Figure 2:**
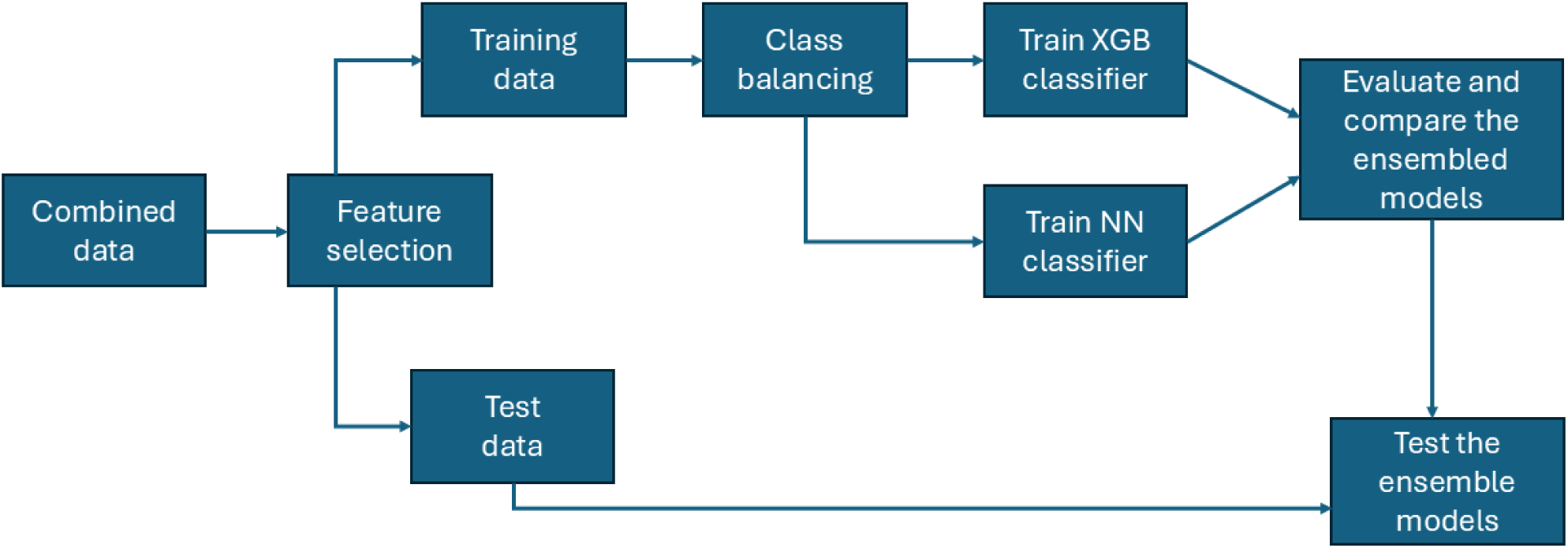
Model training, evaluation and testing.

**Figure 3:**
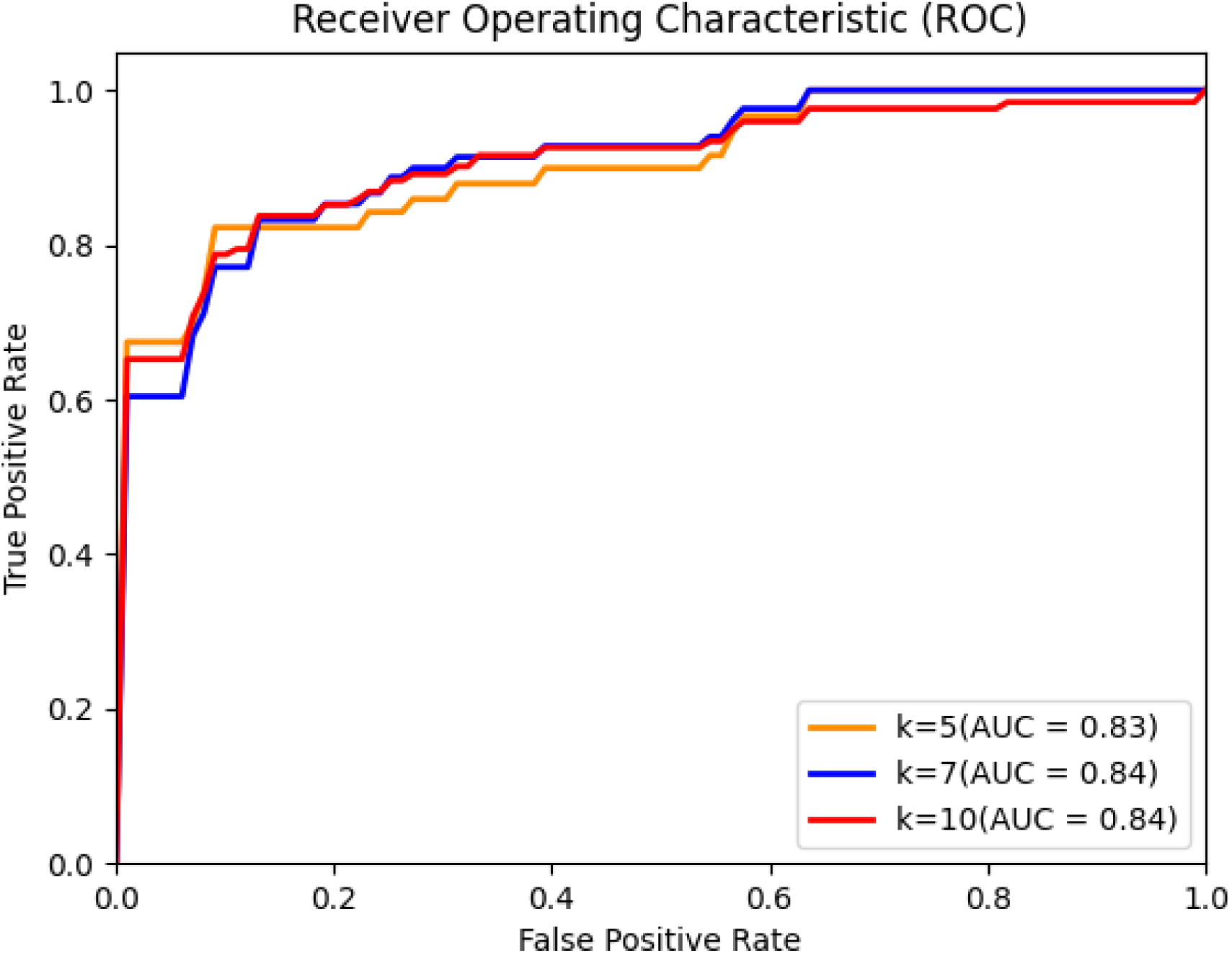
ROC curve of model trained with clinical, deep learning and radiomics features.

The top 10 factors for predicting the OS for each model were added to the appendix. For the combined model, the top 10 features associated with the prediction of the OS were WHOPS, DL_202, DL_301, DL_144, DL_418, DL_25, DL_20, DL_87, DL_229 and DL_133. Except for WHOPS, the rest of these top 10 features are deep learning features. There were no radiomics features in this list. The top 50 features of the combined model included WHOPS, one radiomics feature (original_shape_MajorAxisLength) and 48 deep learning features. WHOPS emerged as the variable with the highest importance in the combined model. Also, in the other three models, WHOPS emerged as the variable with the highest importance.

### Outcome of statistical analysis

We used the Friedman test to analyze the significance of the difference in performance metrics across validation folds across all values of K. When the results of this test indicate a statistically significant difference for the metric, we performed pairwise Wilcoxon signed-rank tests with corrections for multiple comparisons using the FDR method to determine which models had statistically different performances.

The results of the Friedman test indicate a statistically significant difference in the accuracy of the four models with a Chi-squared value of 15.25 and a p-value of 0.0016. This means that there is a statistically significant difference in accuracy for at least one of the models. The outcomes of the pairwise Wilcoxon signed-rank tests are presented in Table 4. There is a statistically significant difference (after FDR correction) between the model trained with clinical features only and the model trained with the combination of all features (p-value = 0.015). Also, there is a statistically significant difference between the model trained with clinical and radiomics features and the model trained with the combination of all features (p-value = 0.004). The Friedman test also indicated statistically significant differences for Precision (Chi-squared: 9.44, p-value: 0.0239), Recall (Chi-squared: 8.94, p-value: 0.0299), F1 score (Chi-squared: 16.85, p-value: 0.0007), AUC (Chi-squared: 17.68, p-value: 0.0005) and MCC (Chi-squared: 17.68, p-value: 0.0005). The Friedman test did not indicate a statistically significant difference for Specificity (Chi-squared: 3.74, p-value: 0.2903). The outcomes of the pairwise Wilcoxon signed-rank tests for Precision, Recall, F1 score, AUC and MCC are added to the Appendix. Similar to accuracy, there is a statistically significant difference (after FDR correction) between the model trained with clinical features only and the model trained with the combination of all features for Recall, F1 score, AUC and MCC. There are also statistically significant differences between the model trained with the combination of clinical and radiomics features and the model trained with the combination of all features for F1 score, AUC and MCC.

**Table 4:**
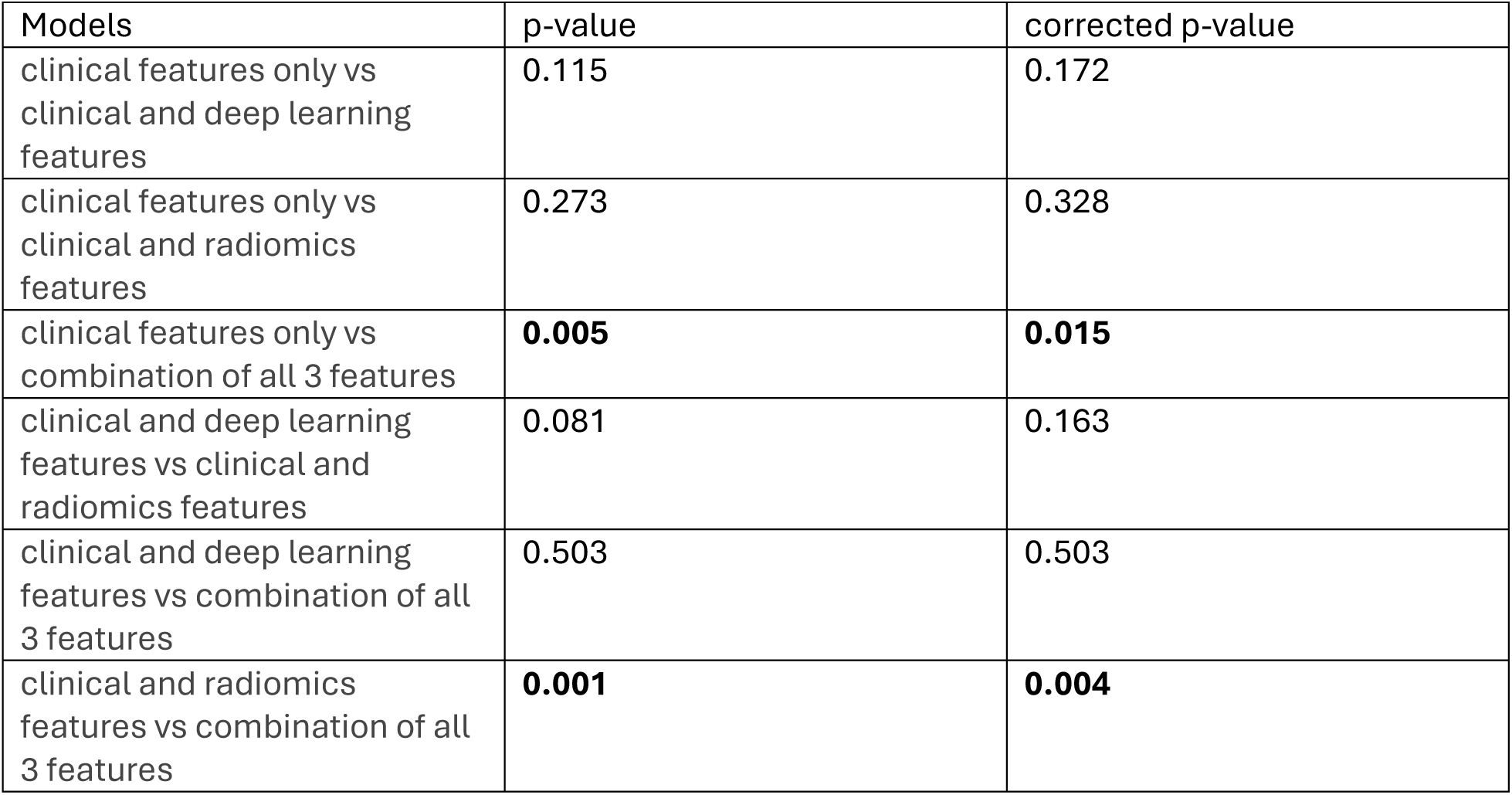
Outcome of statistical analysis for accuracy. In bold are the statistically significant values.

| Models | p-value | corrected p-value |
| --- | --- | --- |
| clinical features only vs clinical and deep learning features | 0.115 | 0.172 |
| clinical features only vs clinical and radiomics features | 0.273 | 0.328 |
| clinical features only vs combination of all 3 features | **0.005** | **0.015** |
| clinical and deep learning features vs clinical and radiomics features | 0.081 | 0.163 |
| clinical and deep learning features vs combination of all 3 features | 0.503 | 0.503 |
| clinical and radiomics features vs combination of all 3 features | **0.001** | **0.004** |

## Discussion

Accurate prediction of 12-month OS in patients with NSCLC enables more personalized and effective therapeutic interventions, ultimately leading to improved treatment outcomes. In this study, we developed and evaluated four distinct machine learning models using various combinations of clinical features (including radiation dose), hand-crafted radiomics features, and deep learning features to predict the 12-month OS in NSCLC patients treated with radiotherapy. The models included: (1) clinical features only; (2) combination of clinical and deep learning features; (3) combination of clinical and radiomics features; and (4) combination of clinical, radiomics, and deep learning features.

Among the four models evaluated, the model incorporating all three feature types (clinical, radiomics, and deep learning features) achieved the highest predictive performance, with an AUC of 0.84 and an accuracy of 85.71%. This model significantly outperformed the model trained solely on clinical features and the model trained with the combination of clinical and radiomics features. Notably, the addition of deep learning features or radiomics alone to the clinical features did not significantly improve the prediction accuracy. This suggests that radiomics and deep learning features may offer complementary prognostic information and that their integration with clinical data yields a synergistic effect, enhancing the model’s predictive capability.

Our findings align with and expand the conclusions of previous studies. Braghetto et al. (2022) combined clinical, hand-crafted radiomics and deep learning features to predict OS for NSCLC patients treated with radiotherapy or chemoradiotherapy [37]. Their study showed that the models combining clinical, radiomics, and deep learning features provided an AUC of 0.67 compared to an AUC of 0.59 using only clinical features. The study of Liao et al. (2024) showed that the combined model achieved an AUC of 0.80 in predicting PFS for NSCLC patients treated with immunotherapy [38]. Our model trained with clinical, deep learning, and radiomics features provided an AUC of 0.84 for predicting OS in NSCLC patients treated with radiotherapy, indicating a potential performance gain. This suggests that methodological enhancements such as applying SMOTE, imputing missing values, and using an ensemble model may contribute to an increase in the prediction performance. Our previous work on the prediction of local control of brain metastases also demonstrated that models integrating clinical, radiomics, and deep learning features outperform those using only a subset of these features [30], suggesting robustness and generalizability of this integrative modeling approach.

Feature importance analysis revealed that WHOPS, a scale used to assess a cancer patient’s general well-being and ability to perform daily activities, was the most important feature in our models, underscoring its known prognostic value of performance status in NSCLC patients. This aligns with the findings from the clinical study of Kawaguchi et al. [39], who identified performance status as an independent prognostic factor for OS in NSCLC. The prognostic strength of WHOPS may be attributed to the fact that the WHOPS reflects the patient’s baseline physical condition, which can influence treatment intensity and decision-making which also carries the risk of confounding our analysis. Differences in baseline status may partially explain the observed variation in survival outcomes. Patients with better performance status and functional capacity tend to have less difficulty in tolerating rigorous cancer treatments and have more favorable outcomes than patients with a worse performance status [61]. Interestingly, the top 10 most predictive features in the combined model included WHOPS and nine deep learning-derived features, but no radiomics features. The top 50 features included WHOPS, one radiomics feature (original_shape_MajorAxisLength) and 48 deep learning features. The planned treatment dose extracted from the RTDOSE DICOM files was also not part of the top 50 features. This could be due to the narrow dose range (54–66 Gy), resulting in low variability across patients and thus offering limited discriminatory power. The higher performance of the combined model when compared to the model trained with clinical and deep learning features suggest that original_shape_MajorAxisLength contains a distinct information that is not captured by clinical or deep learning features. The original_shape_MajorAxisLength feature indicates the degree of elongation of the segmented tumor volume, which can be suggestive of a locally advanced disease. This feature may synergistically interact with the deep learning features and shift decision boundaries of the model in meaningful ways. While the improvement in performance of the combined model was modest when compared to the clinical and deep learning model for the cross validation data sets, the radiomics feature had a greater impact on the performance on the hold-out test set. This shows that the addition of radiomics features to the model improves its robustness and generalization. In general, results suggest that deep learning features may capture more nuanced prognostic information from imaging data than hand-crafted radiomic features. This is in contrast with some other studies which found that radiomics features are more predictive than deep learning features [30, 37]. Further research is needed to evaluate the conditions under which each feature type provides superior prognostic value.

Clinically, these findings have important implications. For patients with unresectable, locally advanced NSCLC, reliably estimating the 12-month OS prior to radiotherapy offers clinicians individualized prognostic information. Such prediction can guide clinical decision-making based on patient-specific risk profiles, enabling the exploration of alternative treatment strategies—for example, implementing more aggressive chemotherapy, immunotherapy, or consolidation therapies in patients with poor predicted prognosis, or considering novel treatment regimens on top of standard care.

Personalized OS prediction before treatment provides a valuable resource for clinicians and patients, supporting shared decision-making and developing personalized treatment plans based on each patient’s unique needs and risk profile.

Although the primary focus of this study was creating a model for predicting the 12-month OS after radiotherapy, the modeling framework can be adapted to other clinical endpoints such as progression-free survival or to different treatment modalities including chemoradiotherapy and chemotherapy. Similarly, this approach can be extended to predicting the OS at other time points, broadening its potential clinical utility.

While our results are promising, this study has limitations. One limitation of this study lies in the tumor segmentation procedure. Since this study included only patients who received definitive radiotherapy, the tumor segmentations were manually delineated by expert radiation oncologists at Maastro clinic, ensuring high precision in defining the ROI. However, this approach also limits the scalability and applicability of the model. Implementing fully automated deep learning–based segmentation methods may improve both efficiency and reproducibility in future studies. Furthermore, for a more rigorous evaluation of the efficacy and versatility of the models, further investigations involving larger patient cohorts with multi-institutional data are recommended. Our results are from a small cohort of radiotherapy patients from a single institution. An external validation dataset could significantly improve the generalizability of the OS prediction model and enrich confidence in its broader applicability across diverse patient populations and healthcare domains. Also, because our analysis is based on historical data the model only provides prognostic information and cannot be used to support treatment selection such as between concurrent or sequential radiotherapy and other therapies.

In conclusion, our results demonstrate that integrating clinical, radiomics, and deep learning features substantially improves OS prediction in NSCLC patients treated with radiotherapy. The developed model provides a valuable tool for risk stratification, treatment planning, and patient counseling, paving the way for more personalized and more effective cancer care.

## Acknowledgements

We would like to thank Dr. Dirk De Ruysscher (Department of Radiation Oncology, Maastro) and Dr. Lizza Hendriks (Department of Pulmonary Diseases, Maastricht University Medical Centre+) for their valuable contribution in providing clinical data for this study.

## Data Availability

The data used for this study is available at Maastro and is accessible upon request after necessary approvals.

## Consent for publication

Not applicable.

## Contributions

All authors contributed to this research.

## Ethics approval

This study is part of the AI in Medical Imaging for novel Cancer User Support (AMICUS) project. This study was approved by the Institutional Review Board of Maastro Clinic (P0617) and Ethics Committee of Maastricht University Medical Centre (METC 2023-0377). This study adhered to the ethical principles outlined in the Declaration of Helsinki.

## Clinical trial number

Not applicable.

## List of abbreviations

OS: Overall Survival
NSCLC: Non-Small Cell Lung Cancer
DL: Deep Learning
AUC: Area Under the Receiver Operating Characteristic Curve
SCLC: Small Cell Lung Cancer
PFS: Progression-Free Survival
HU: Hounsfield Unit
HFS: Head-First Supine
WHOPS: World Health Organisation Performance Status
CKD-EPI: Chronic Kidney Disease Epidemiology Collaboration
CRP: C-reactive protein
FEV₁: Forced Expiratory Volume in 1 second
DLCO: Diffusing Capacity of the Lungs for Carbon Monoxide
Gy: Gray
ROI: Region Of Interest
GLCM: Gray Level Cooccurrence Matrix
GLDM: Gray Level Dependence Matrix
GLRLM: Gray Level Run Length Matrix
GLSZM: Gray Level Size Zone Matrix
NGTDM: Neighbouring Gray Tone Difference Matrix
RFE: Recursive Feature Elimination
XGB: XGBoost
NN: Neural Network
ReLU: Rectified Linear Unit
MCC: Matthews Correlation Coefficient
ROC: Receiver Operating Characteristic
FDR: False Discovery Rate

## Appendix

**Full list of Radiomics features**

Full list of Radiomics features:

original_shape_Elongation

original_shape_Flatness

original_shape_LeastAxisLength

original_shape_MajorAxisLength

original_shape_Maximum2DDiameterColumn

original_shape_Maximum2DDiameterRow

original_shape_Maximum2DDiameterSlice

original_shape_Maximum3DDiameter

original_shape_MeshVolume

original_shape_MinorAxisLength

original_shape_Sphericity

original_shape_SurfaceArea

original_shape_SurfaceVolumeRatio

original_shape_VoxelVolume

original_firstorder_10Percentile

original_firstorder_90Percentile

original_firstorder_Energy

original_firstorder_Entropy

original_firstorder_InterquartileRange

original_firstorder_Kurtosis

original_firstorder_Maximum

original_firstorder_MeanAbsoluteDeviation

original_firstorder_Mean

original_firstorder_Median

original_firstorder_Minimum

original_firstorder_Range

original_firstorder_RobustMeanAbsoluteDeviation

original_firstorder_RootMeanSquared

original_firstorder_Skewness

original_firstorder_TotalEnergy

original_firstorder_Uniformity

original_firstorder_Variance

original_glcm_Autocorrelation

original_glcm_ClusterProminence

original_glcm_ClusterShade

original_glcm_ClusterTendency

original_glcm_Contrast

original_glcm_Correlation

original_glcm_DifferenceAverage

original_glcm_DifferenceEntropy

original_glcm_DifferenceVariance

original_glcm_Id

original_glcm_Idm

original_glcm_Idmn

original_glcm_Idn

original_glcm_Imc1

original_glcm_Imc2

original_glcm_InverseVariance

original_glcm_JointAverage

original_glcm_JointEnergy

original_glcm_JointEntropy

original_glcm_MCC

original_glcm_MaximumProbability

original_glcm_SumAverage

original_glcm_SumEntropy

original_glcm_SumSquares

original_gldm_DependenceEntropy

original_gldm_DependenceNonUniformity

original_gldm_DependenceNonUniformityNormalized

original_gldm_DependenceVariance

original_gldm_GrayLevelNonUniformity

original_gldm_GrayLevelVariance

original_gldm_HighGrayLevelEmphasis

original_gldm_LargeDependenceEmphasis

original_gldm_LargeDependenceHighGrayLevelEmphasis

original_gldm_LargeDependenceLowGrayLevelEmphasis

original_gldm_LowGrayLevelEmphasis

original_gldm_SmallDependenceEmphasis

original_gldm_SmallDependenceHighGrayLevelEmphasis

original_gldm_SmallDependenceLowGrayLevelEmphasis

original_glrlm_GrayLevelNonUniformity

original_glrlm_GrayLevelNonUniformityNormalized

original_glrlm_GrayLevelVariance

original_glrlm_HighGrayLevelRunEmphasis

original_glrlm_LongRunEmphasis

original_glrlm_LongRunHighGrayLevelEmphasis

original_glrlm_LongRunLowGrayLevelEmphasis

original_glrlm_LowGrayLevelRunEmphasis

original_glrlm_RunEntropy

original_glrlm_RunLengthNonUniformity

original_glrlm_RunLengthNonUniformityNormalized

original_glrlm_RunPercentage

original_glrlm_RunVariance

original_glrlm_ShortRunEmphasis

original_glrlm_ShortRunHighGrayLevelEmphasis

original_glrlm_ShortRunLowGrayLevelEmphasis

original_glszm_GrayLevelNonUniformity

original_glszm_GrayLevelNonUniformityNormalized

original_glszm_GrayLevelVariance

original_glszm_HighGrayLevelZoneEmphasis

original_glszm_LargeAreaEmphasis

original_glszm_LargeAreaHighGrayLevelEmphasis

original_glszm_LargeAreaLowGrayLevelEmphasis

original_glszm_LowGrayLevelZoneEmphasis

original_glszm_SizeZoneNonUniformity

original_glszm_SizeZoneNonUniformityNormalized

original_glszm_SmallAreaEmphasis

original_glszm_SmallAreaHighGrayLevelEmphasis

original_glszm_SmallAreaLowGrayLevelEmphasis

original_glszm_ZoneEntropy

original_glszm_ZonePercentage

original_glszm_ZoneVariance

original_ngtdm_Busyness

original_ngtdm_Coarseness

original_ngtdm_Complexity

original_ngtdm_Contrast

original_ngtdm_Strength

**Performance of XBG classifier for test data set:**

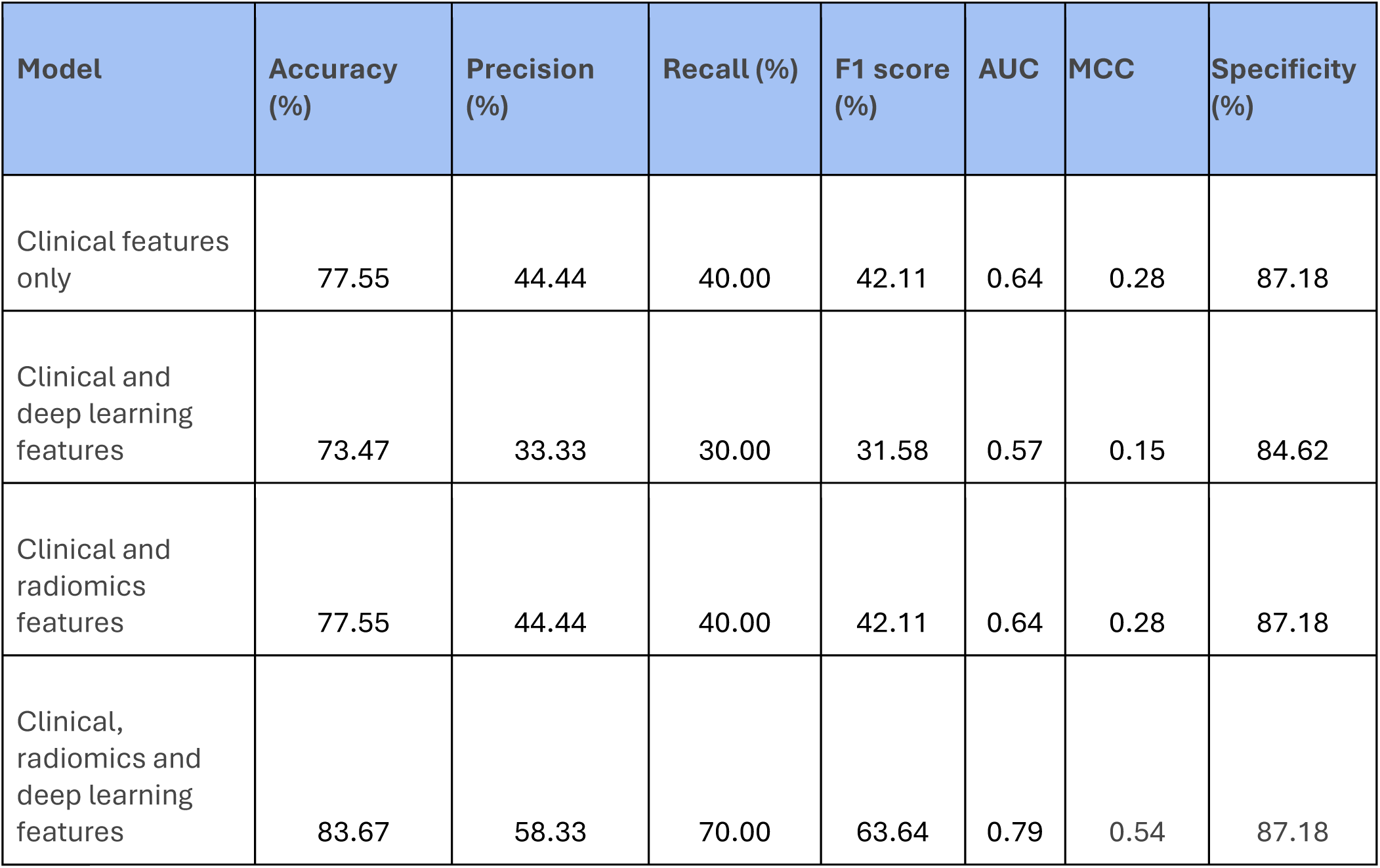

| **Model** | **Accuracy (%)** | **Precision (%)** | **Recall (%)** | **F1 score (%)** | **AUC** | **MCC** | **Specificity (%)** |
| --- | --- | --- | --- | --- | --- | --- | --- |
| Clinical features only | 77.55 | 44.44 | 40.00 | 42.11 | 0.64 | 0.28 | 87.18 |
| Clinical and deep learning features | 73.47 | 33.33 | 30.00 | 31.58 | 0.57 | 0.15 | 84.62 |
| Clinical and radiomics features | 77.55 | 44.44 | 40.00 | 42.11 | 0.64 | 0.28 | 87.18 |
| Clinical, radiomics and deep learning features | 83.67 | 58.33 | 70.00 | 63.64 | 0.79 | 0.54 | 87.18 |

**Performance of NN classifier for test data set:**

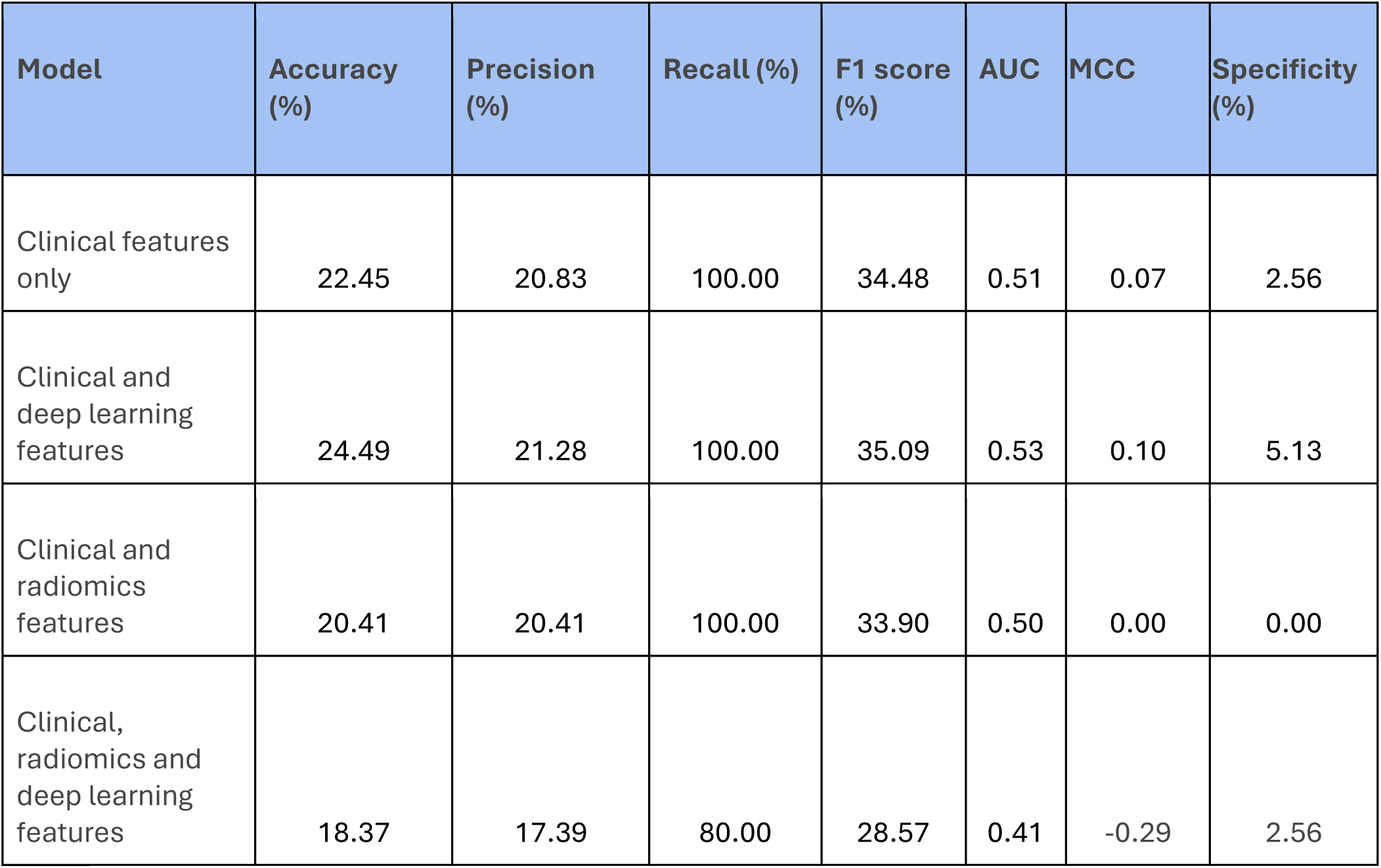

**Outcome of statistical analysis for Precision:**

| Models | p-value | corrected p-value |
| --- | --- | --- |
| clinical features only vs clinical and deep learning features | 0.5897 | 0.6885 |
| clinical features only vs clinical and radiomics features | 0.0583 | 0.1522 |
| clinical features only vs combination of all 3 features | 0.1392 | 0.2088 |
| clinical and deep learning features vs clinical and radiomics features | 0.0761 | 0.1522 |
| clinical and deep learning features vs combination of all 3 features | 0.6885 | 0.6885 |
| clinical and radiomics features vs combination of all 3 features | **0.0005** | **0.0028** |

**Outcome of statistical analysis for Recall:**

| Models | p-value | corrected p-value |
| --- | --- | --- |
| clinical features only vs clinical and deep learning features | **0.0084** | **0.0252** |
| clinical features only vs clinical and radiomics features | 0.9195 | 0.9642 |
| clinical features only vs combination of all 3 features | **0.0018** | **0.0108** |
| clinical and deep learning features vs clinical and radiomics features | 0.1013 | 0.1520 |
| clinical and deep learning features vs combination of all 3 features | 0.9642 | 0.9642 |
| clinical and radiomics features vs combination of all 3 features | 0.0379 | 0.0757 |

**Outcome of statistical analysis for F1 score:**

| Models | p-value | corrected p-value |
| --- | --- | --- |
| clinical features only vs clinical and deep learning features | 0.1585 | 0.2378 |
| clinical features only vs clinical and radiomics features | 0.3750 | 0.4500 |
| clinical features only vs combination of all 3 features | **0.0014** | **0.0043** |
| clinical and deep learning features vs clinical and radiomics features | 0.0637 | 0.1273 |
| clinical and deep learning features vs combination of all 3 features | 0.6327 | 0.6327 |
| clinical and radiomics features vs combination of all 3 features | **0.0002** | **0.0013** |

**Outcome of statistical analysis for AUC:**

| Models | p-value | corrected p-value |
| --- | --- | --- |
| clinical features only vs clinical and deep learning features | 0.1585 | 0.2378 |
| clinical features only vs clinical and radiomics features | 0.3536 | 0.4244 |
| clinical features only vs combination of all 3 features | **0.0014** | **0.0042** |
| clinical and deep learning features vs clinical and radiomics features | 0.0581 | 0.1162 |
| clinical and deep learning features vs combination of all 3 features | 0.7024 | 0.7024 |
| clinical and radiomics features vs combination of all 3 features | **0.0001** | **0.0008** |

**Outcome of statistical analysis for MCC:**

| Models | p-value | corrected p-value |
| --- | --- | --- |
| clinical features only vs clinical and deep learning features | 0.1917 | 0.2875 |
| clinical features only vs clinical and radiomics features | 0.3536 | 0.4244 |
| clinical features only vs combination of all 3 features | **0.0014** | **0.0043** |
| clinical and deep learning features vs clinical and radiomics features | 0.0581 | 0.1162 |
| clinical and deep learning features vs combination of all 3 features | 0.6327 | 0.6327 |
| clinical and radiomics features vs combination of all 3 features | **0.0001** | **0.0008** |

**Top 10 features:**

The top 10 features of the clinical only model are WHOPS, NSCLC stage, laterality, N classification, presence of cerebra vascular issues, durvalumab administered indicator, CRP (C-reactive protein) level, creatinine level, presence of cardiac surgery and presence of vascular issues.

The top 10 features of clinical with deep learning model are WHOPS, DL_426, DL_98, DL_202, DL_288, DL_384, DL_30, DL_414, DL_301 and DL_122.

The top 10 features of clinical with radiomics model are WHOPS, NSCLC stage, presence of cardiac surgery, presence of cerebra vascular issues, creatinine level, N classification, CRP (C-reactive protein) level, laterality, radiotherapy treatment dose, and original_shape_MinorAxisLength.

**ROC Curves:**

**Figure 4:**
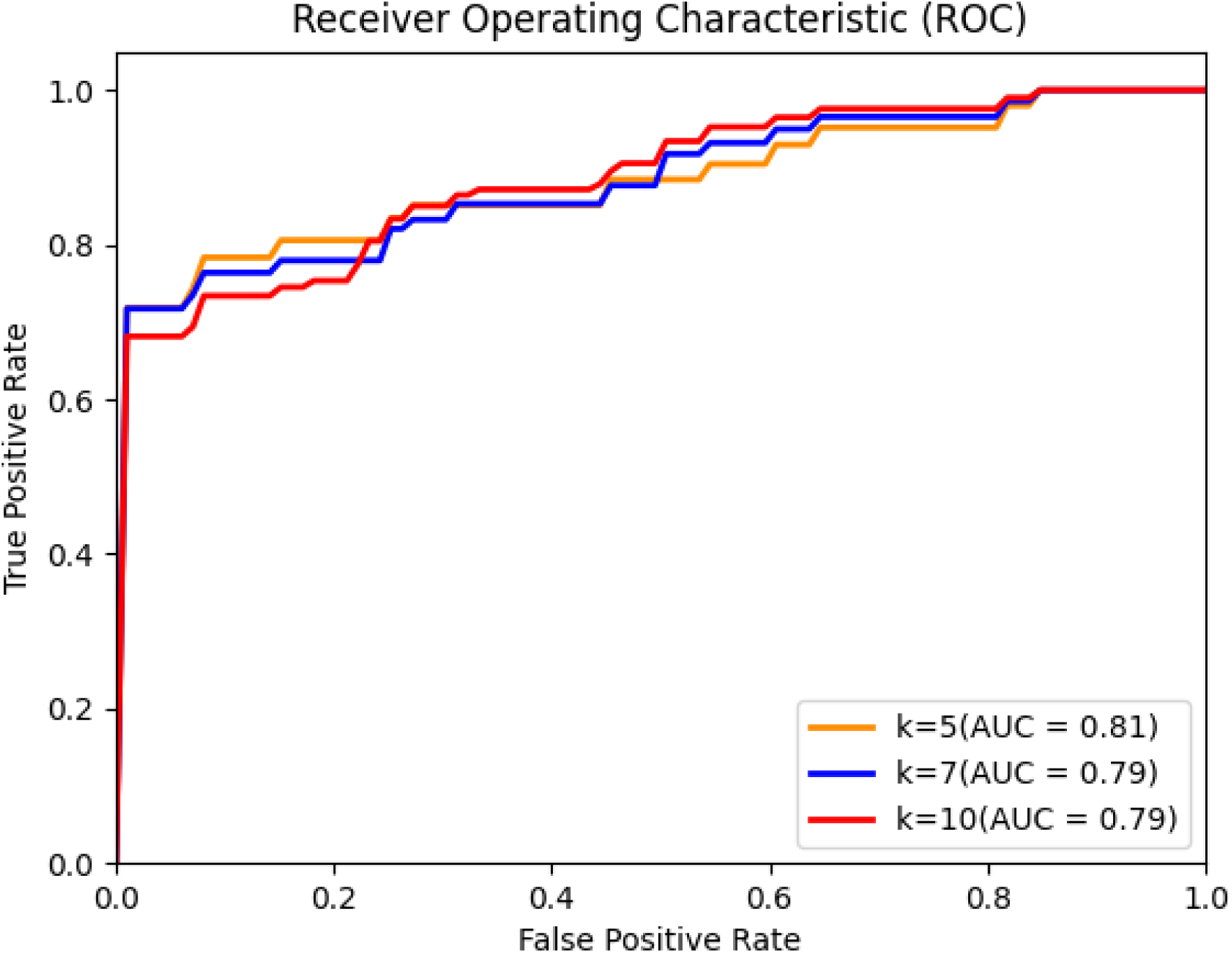
ROC curve of model trained with clinical features only.

**Figure 5:**
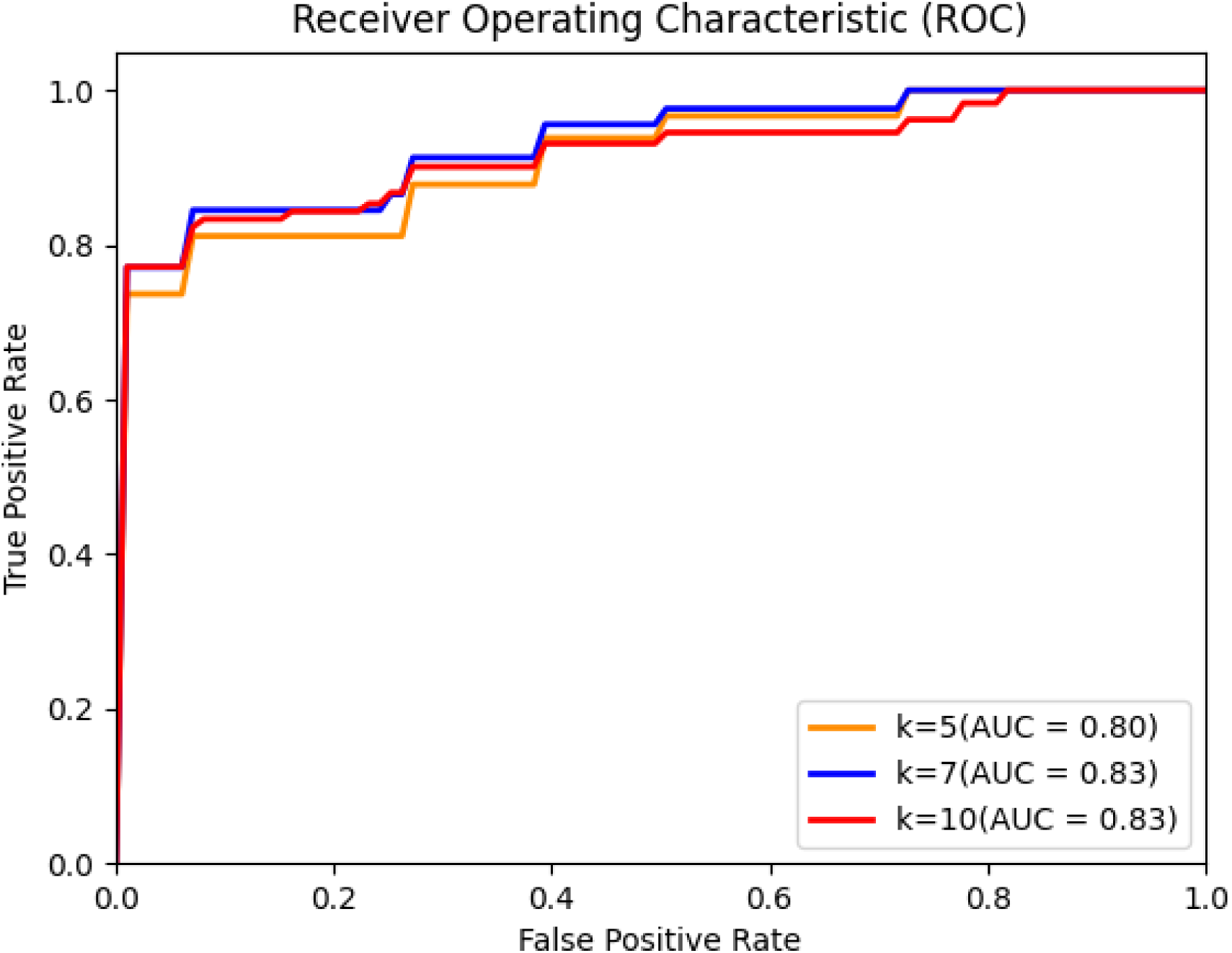
ROC curve of model trained with clinical and deep learning features.

**Figure 6:**
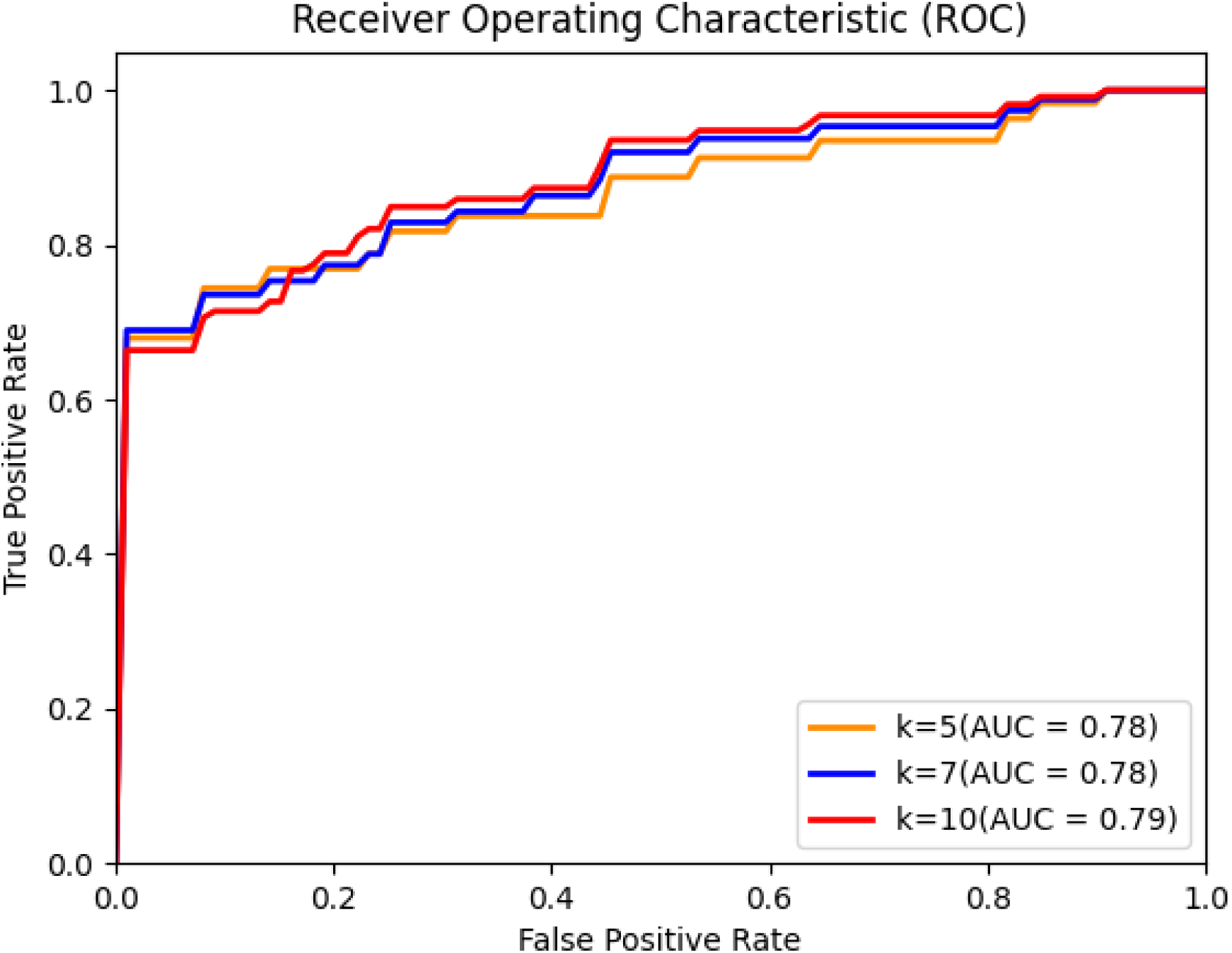
ROC curve of model trained with clinical and radiomics features.

